# Autism Prevalence in Chile: Unmet Special Education Needs using Data Linkage and Bayesian Analysis of Three Million School-Aged Children

**DOI:** 10.1101/2024.06.25.24309483

**Authors:** Andres Roman-Urrestarazu, Adele Tyson, Gabriel Gatica-Bahamonde, Robin van Kessel, Justin Yang, Carola Mansilla, Isabel Zuniga, Alejanda Mendez-Fadol, Blanca Larrain, Ricardo Garcia, Damaris Koch, Wim Groot, Milena Pavlova, Katarzyna Czabanowska, Tamsin Ford

**Author notes:** **Corresponding author:** Dr Andres Roman-Urrestarazu Department of Psychiatry University of Cambridge Douglas House, 18b Trumpington Road Cambridge, CB2 8AH. These authors contributed equally (Andres Roman-Urrestarazu, Adele Tyson).

## Abstract

Prevalence estimates of autism spectrum disorder (henceforth autism) in Latin America thus far have been limited by a lack of reliable population-level data. We analyzed autism school prevalence across 29 Chilean health service regions for students aged 6–18 years, standardized by age and sex. We validated these results using electronic health records from one of Chile’s largest regional health service, the Servicio de Salud Araucania Sur (SSAS). We then projected Bayesian prevalences, reporting nationally, and by health service, ethnicity, immigration background, and rurality. We found a standardized national school autism prevalence of 0.46% (95% CI, 0.46%-0.47%), with boys having six times higher odds of autism than girls (OR 6.10 [95%CI: 5.82–6.41]). The sex - and age-adjusted clinical prevalence in the SSAS trust was 1.22% (95% CI: 1.16%-1.28%) and the projected Bayesian national autism prevalence was 1.31% (95% Credible Interval: 1.25%-1.38%). Our results indicate a higher autism prevalence than previously reported in the south of the Araucania region with observed disparities in prevalence across sex, ethnic groups, and health services.

## 1. Main

Autism spectrum disorder (henceforth autism) is a neurodevelopmental condition that affects social interaction and communication and affects 1–2% of the global population^1,2^. Recently, there has been a growing interest in understanding autism prevalence, particularly using large research designs such as school registries and access to special educational needs (SEN) services from school registry data^1,2^. However, little is known about how this relates to clinical prevalence, and few studies have linked national registries to electronic health records^3^ to study the gap between those diagnosed and those receiving support at school^4–6^. One challenge of studying autism prevalence in Latin America and the Caribbean is the lack of reliable data sources^4,5^, reinforced by only 2.3% of global published prevalence studies to date being based in this region^4,5,7^, as well as only one new autism prevalence study identified in Latin America, in Ecuador specifically, since 2012^4^. Chile is a high income country with a population of 19 million people^8^ and ranks as one of the wealthiest and most socioeconomically unequal Latin American countries^9^, with significant disparities in health outcomes according to socioeconomic status^10,11^. Inequities in access to care, higher unmet needs among those unable to pay, and longer waiting times for those using non-private providers, affect autistic people^6,12^. In 2023, to address inequalities in autism diagnosis and screening, Chile legislated a new autism law (Law N° 21.545)^13^ which establishes inclusion, comprehensive care, and the need for accurate epidemiological estimates in order for this law to have a meaningful effect.

As such, we aimed to examine the prevalence of autism in Chilean school-aged children aged 6 to 18 years linking registry data from the Chilean school SEN inclusion program (Programa de Integracion Escolar; PIE) to electronic health records from one of Chile’s 29 regional health services, the Servicio de Salud Araucania Sur (SSAS). Specifically, we (a) investigated the prevalence of autistic children in schools using the Chilean PIE, (b) assessed access determinants to autism school SEN services in Chile using a two-level mixed effects logistic regression model, (c) explored differences and service gaps between school-level autism and clinical service prevalence by assessing the difference in prevalence estimates between the school registry and clinical records using a probabilistic linkage of electronic health records in the SSAS service, (d) estimated the unmet need for SEN services based on disparities between clinical diagnosis and school-level services, and (e) made a national prevalence estimation based on the findings of our SSAS clinical record analysis, which were extrapolated to Chile’s remaining 28 health services using a Bayesian prevalence model. For the Bayesian prevalence model, we first constructed baseline national and health service autism prevalence rates using only the national school registry dataset, which served as the conservative prevalence inputs for the Bayesian prevalence model in order to calculate the lower limit of autism prevalence in Chile. We then linked school registry data with clinical data to account for potential underestimations in the school registry data, which served as a more generous prevalence input for the Bayesian model to calculate the upper limit of autism prevalence. Finally, we constructed a uniform prior for each respective health service that spanned the full range of possible autism prevalences, from the lowest estimates provided by school registry data to the highest suggested by linked data. This allowed the Bayesian model to explore all plausible values of autism prevalence without bias towards either end of the spectrum. This study has important implications for policy and resource allocation related to the education and support of children with autism in Chile and in Latin America and the Caribbean more broadly.

## 2. Results

### 2.1. Descriptive Statistics and Frequentist Prevalence Estimation using School Data

Our final school dataset consisted of 3,056,306 children aged 6–18 years (boys N=1,569,082; 51.34%). A PIE code was recorded for N=339,968 children (11.12%) and for 48.41% of the 12,077 Chilean schools participating in the PIE program. A total of 14,549 students with autism were identified in the school registry (boys N=12,571 [86.40%], girls N= 1,978 [13.60%]). The adjusted prevalence of autism in the national sample of schools was 0.46% (95%CI: 0.45–0.47%), with a prevalence in boys of 0.79% (95%CI: 0.77–0.80%) and in girls of 0.13% (95%CI: 0.13–0.14%), with a boys-to-girls ratio of 6:1. Further demographic details are shown in Table 1.

**Table 1.**
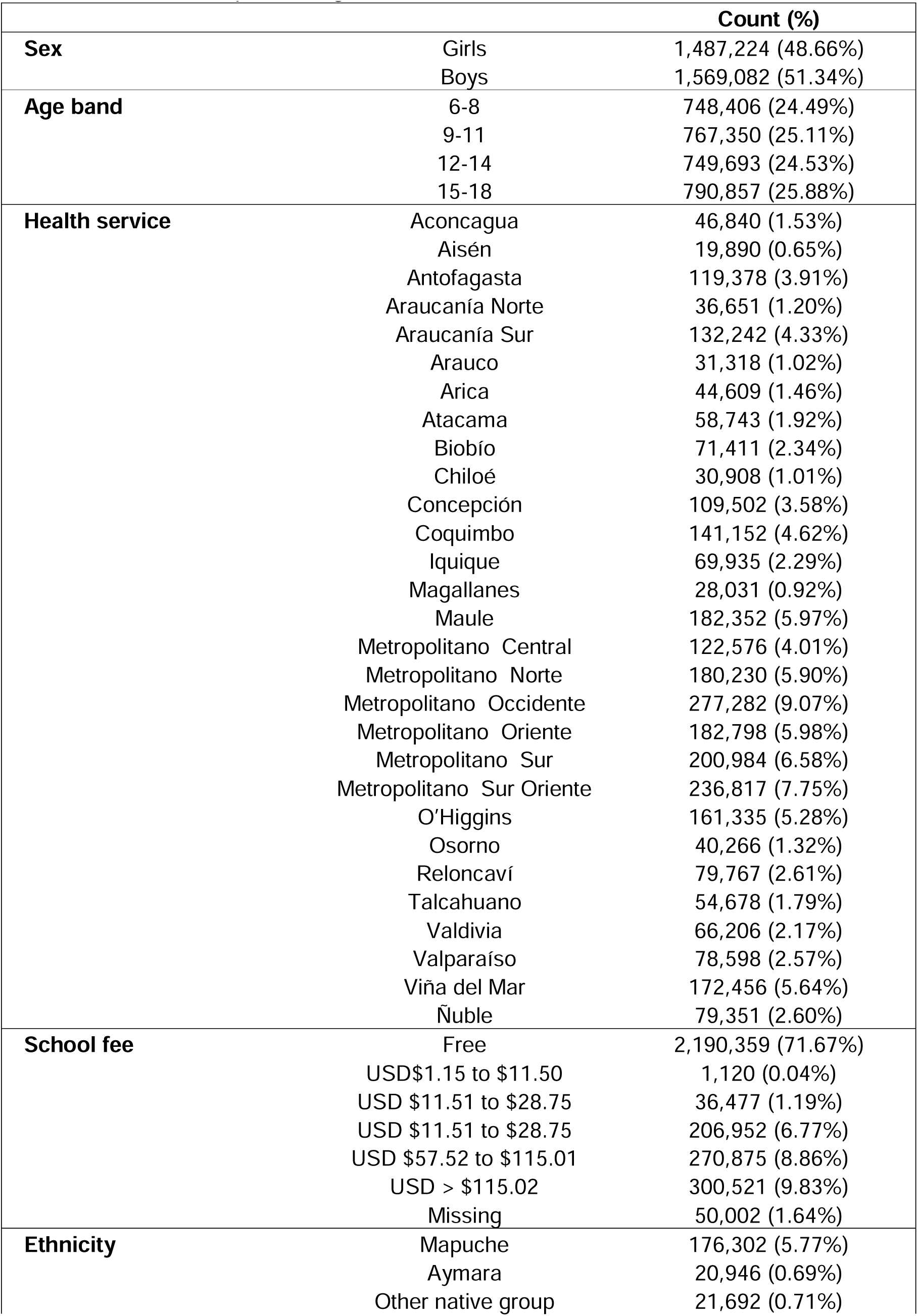
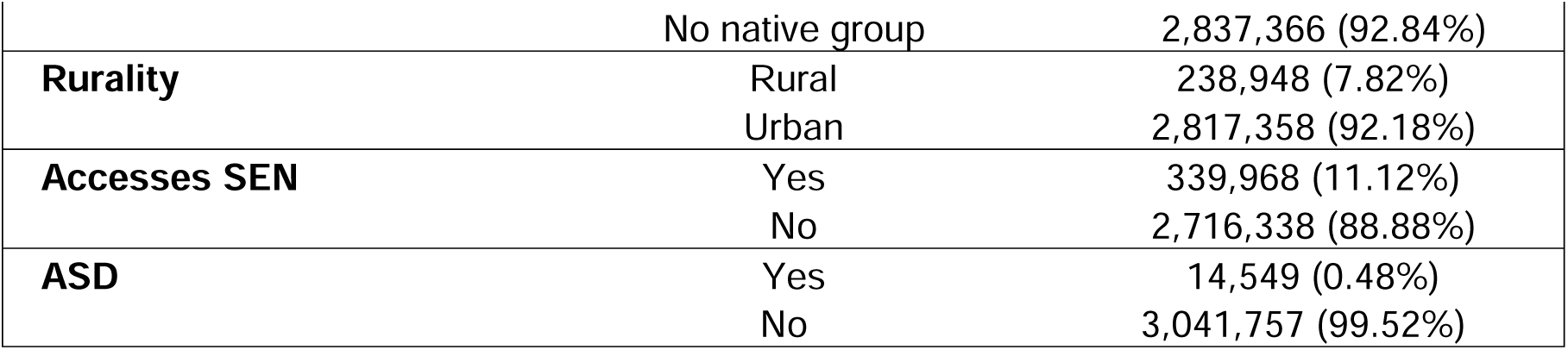
Count and percentage of features’ values in the school dataset.

The sample included N=176,302 (5.77%) Mapuche and N=20,946 (0.69%) Aymara pupils and N=21,692 students declared to belong to other indigenous groups, which were labelled as other due to disclosure risks. We found an adjusted autism school prevalence of 0.35% (0.32%-0.38%) for Mapuche pupils, with 0.07% (95% CI: 0.05%-0.09%) of Mapuche girls and 0.62% (95%CI: 0.57% -0.67%) of Mapuche boys (Male-to-Female Ratio, MFR: 8.86:1) being autistic, and 0.65% (95%CI: 0.55% -0.80%) for Aymara pupils with a prevalence of 0.17% (95%CI: 0.17%-0.24%) for Aymara girls and 1.15% for Aymara boys (MFR: 6.8:1), and 0.47% for all other indigenous groups (95% CI: 0.39%-0.63%), with a prevalence for girls of 0.12% (95%CI: 0.05%0.19%) and for boys of 0.81% (95%CI: 0.63%–0.98%) for all other ethnic indigenous groups (MFR:6.75:1). These rates are lower and higher, respectively, than the rates for those not declaring to belong to these groups (0.47% (95%CI: 0.46%-0.48%)), with an autism prevalence of 0.13% (95%CI: 0.13%-0.14%) for girls and 0.79% (95%CI: 0.78-0.81%) for boys. This contrasts to an MFR in non-indigenous children of 6.1:1. Immigrant children reported an adjusted autism prevalence of 0.19% (95%CI: 0.17%-0.21%). Further details are shown in Table 2.

**Table 2.**
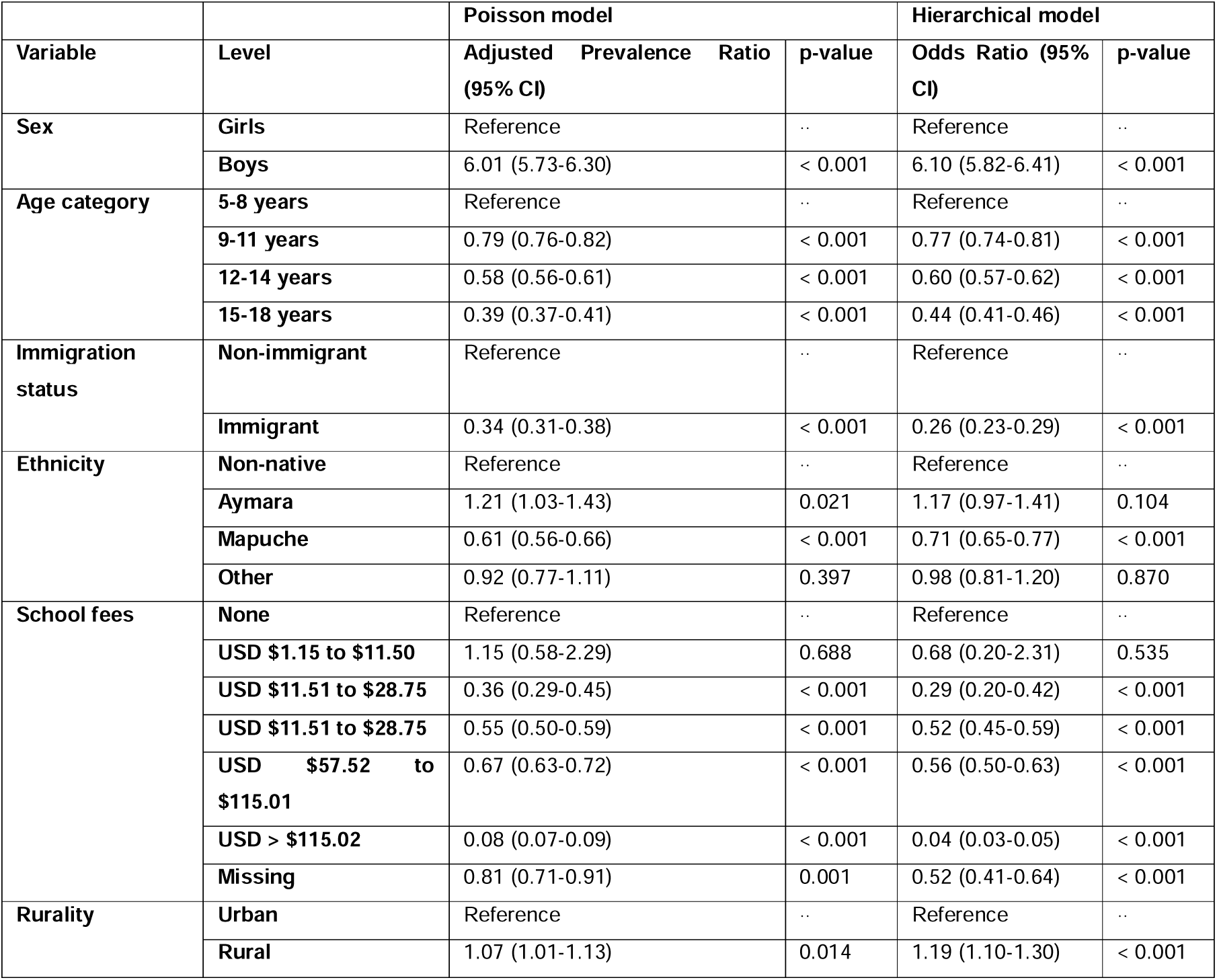
Poisson and hierarchical regression estimates for SEN Access. The hierarchical model is nested at the school and commune levels.

Autism prevalence varied across Chile’s 29 health services (eTable 1), with the highest adjusted prevalence reported in Ñuble (1.29% [95% CI, 1.21%-1.37%]) and the lowest in Metropolitano Norte (0.29% [0.26%-0.31%]). Prevalence was also low in the Metropolitano health services serving Santiago, Chile’s largest city, which all had a prevalence below 0.40%. Autism peaked in the 6–8 age band across all services with the exception of Chiloé and Magallanes, where it peaked in the 9–11 band (eTable 2 in the appendix).

### 2.2. Analysis of Access to Autism SEN in Schools Using Poisson Regression

Our two level (school and commune) mixed effects logistic regression model assessed determinants of SEN access and shows similar results, with boys having six times higher odds of having a diagnosis and of receiving SEN support than girls (OR 6.10 [95%CI: 5.82–6.41]). Compared to children aged 5–8 years, all other age groups showed lower odds of autism, as did immigrants and Mapuche children.

Compared to children who do not pay school fees, all children in school fee categories of $10,001 or higher reported lower odds of autism. Children from rural areas reported higher odds of autism compared to children from urban regions (1.19 [95%CI: 1.10–1.30]). Our likelihood-ratio test asserts that our two level mixed effects model is an improvement over a simple Poisson regression mode (χ^2^ = 9112.46; p < 0.001). Further details are shown in Table 2.

### 2.3. Probabilistic Data Linkage, Unmet SEN Need, and Autism Prevalence from the Clinical Validation Sample

After matching by sex and date of birth, we obtained 293 pairs. Probabilistic matching performed using sex, date of birth, commune of residence, and proxies for socioeconomic status (SES) with selection of possible matches to create a bijective set of matches resulted in 233 matches of unique SSAS school and patient records (see Figure 1). Cohen’s Kappa for inter-rater reliability on the diagnostic validation subsample was 0.97, indicating excellent agreement between electronic health record diagnoses and the diagnoses from our independent clinicians. This corresponds to 47.65% of school records for students with autism in SSAS having a match in SSAS patient records, to 16.93% of patient records having a match in SSAS school records, and 17.07% of unique patients having a match in SSAS school records. Only one match to an SSAS school record was made for each patient who had lived in more than one commune and who, therefore, appeared more than once in the patient data. This means that matching was bijective for SSAS school records and unique patients. After linking SSAS school and patient data, we found 1,132 or 69.9% patients with autism who could not be matched to students in the school registry. This represents the unmet need of SSAS students with autism who did not access school-based support. Combining these additional cases with the 488 or 30.1% students who accessed a Differential Special Education Grant *(Subvención de Educación Especial Diferencial*; SEED) for autism resulted in 1,620 school-aged children with autism in SSAS, which has a total population of N=132,242 children aged 6–18 years.

**Figure 1.**
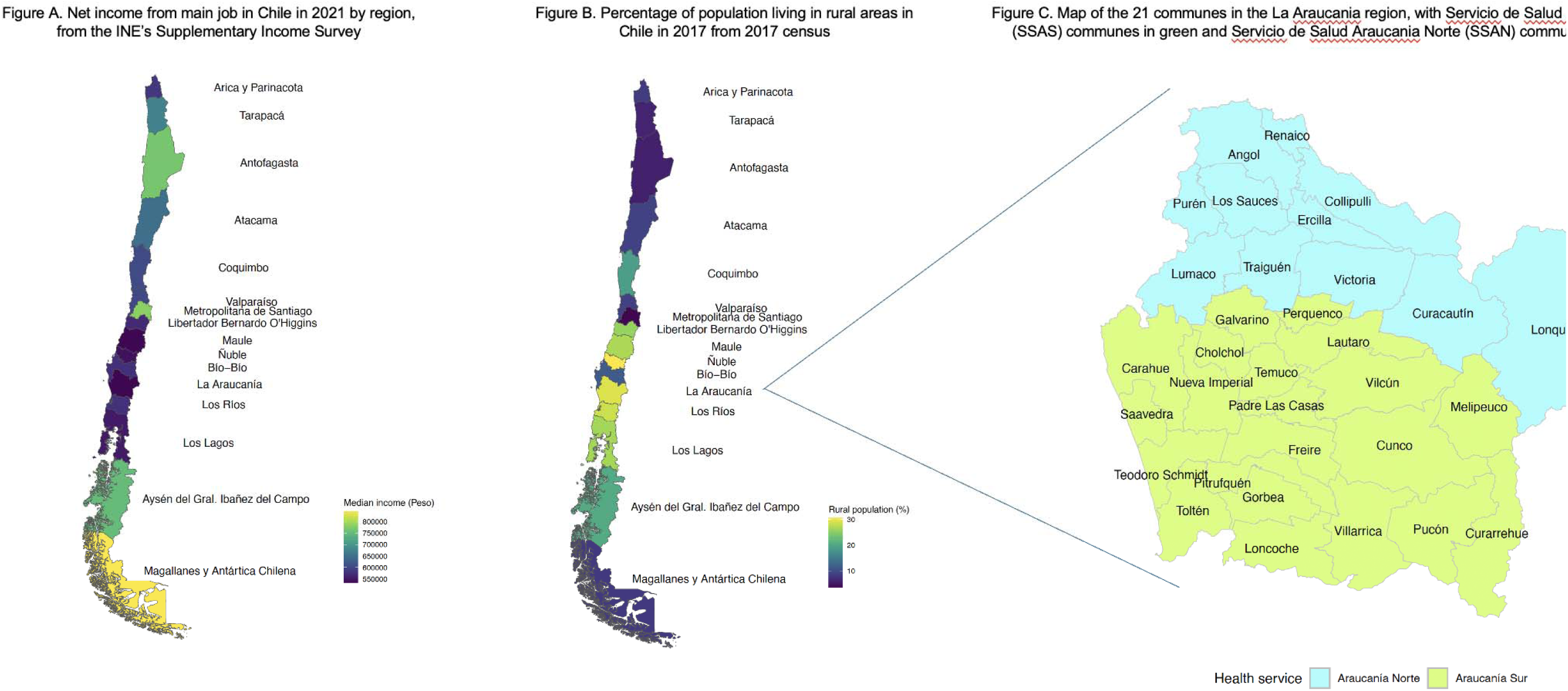
Map of Chile’s regions and rurality with zoom into the Araucania region.

The updated crude prevalence of autism in SSAS was 1.23% (95%CI: 1.17–1.28%) and the updated adjusted prevalence was 1.22% (95%CI: 1.16–1.28%). The adjusted SSAS prevalence for girls was 0.47% (95%CI: 0.41–0.53%) and for boys was 1.95% (95%CI: 1.84–2.06%). This gave an updated male-to-female ratio of 4.18:1 after data linkage, which is smaller than the 6:1 ratio in the school data and shows concerning differences in access to SEN services in girls. Autism prevalence in this clinical sample was highest among children aged 6–8 years and those starting primary school, at 1.54% (95%CI: 1.41–1.68%); this decreased with age (Table 3).

**Table 3:**
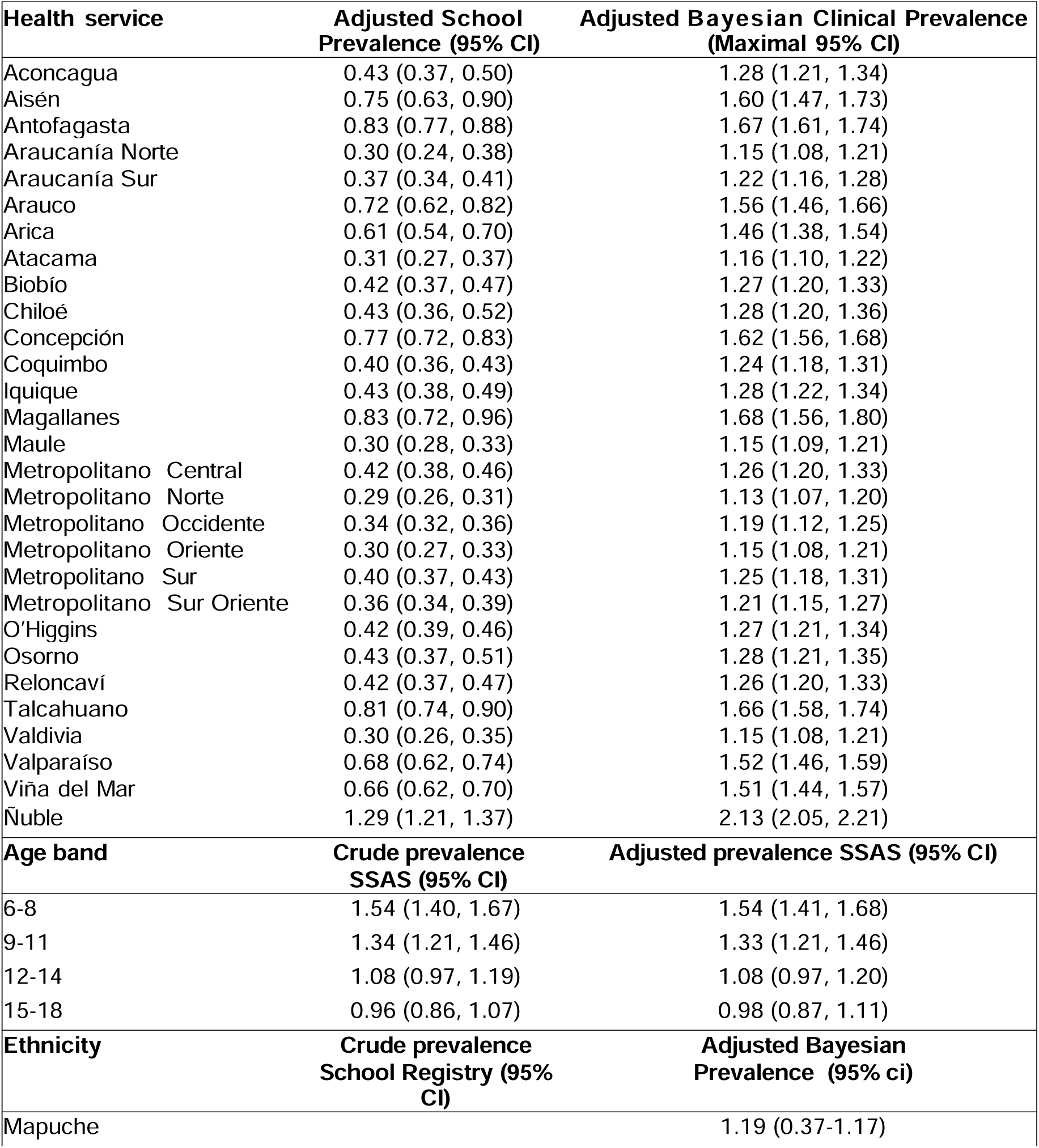
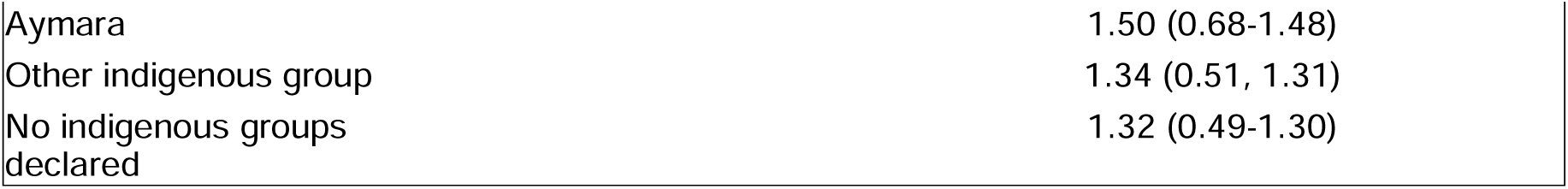
Adjusted prevalence and adjusted updated prevalence of ASD by health service in Chile. Adjusted prevalence is from school data only. Adjusted updated prevalence is from linkage of school data and patient data. Prevalence for Servicio de Salud Araucanía Sur (SSAS) was calculated directly from linkage results. Prevalence for other health services was calculated by adding the adjusted prevalence delta to each health service’s adjusted prevalence from the school data only. Adjusted prevalence has 95% gamma confidence intervals. The width of the adjusted updated prevalence confidence intervals is the maximum of the school data adjusted prevalence confidence intervals for each health service, and the adjusted prevalence delta confidence interval, except for SSAS which has the 95% gamma confidence intervals found earlier.

### 2.4. Bayesian Prevalence Analysis

Bayesian clinical prevalence projections by health service with school level autism prevalence prior probability distribution (priors) are shown in Figure 2, representing the assumed probability distribution of autism prevalence before considering specific evidence from electronic health records. This approach was implemented by the selection of four priors that reflected varying levels of knowledge and assumptions about autism prevalence from our data: conjugate beta priors based on national and health service school data, priors based on linked school registry and clinical data offering higher prevalence estimates, and uniform priors to cover the entire plausible range of values. The model then iteratively updated these priors with actual data inputs to produce posterior distributions, which provided a Bayesian assessment of autism prevalence, taking into account both the known and uncertain elements of the datasets. The posterior prevalence peaks from the school registry can be considered lower bounds for the true autism prevalence in each health service as they are based on children diagnosed with autism who receive SEN support in Chilean schools. Bayesian prevalence projections were pulled upward toward their priors by our approximate delta of unmet need in linked data prior and the uniform prior, and their posterior credible intervals (Table 3). This process produced a national Bayesian autism prevalence of 1.31% (95% Credible Interval [CrI]: 1.25 -1.38%). This, in turn, posits that Chile has 40,113 children aged 6–18 with autism, corresponding to an estimated unmet need of 25,903 children lacking SEN access for autism (posited count and unmet need are age- and sex-adjusted).

**Figure 2.**
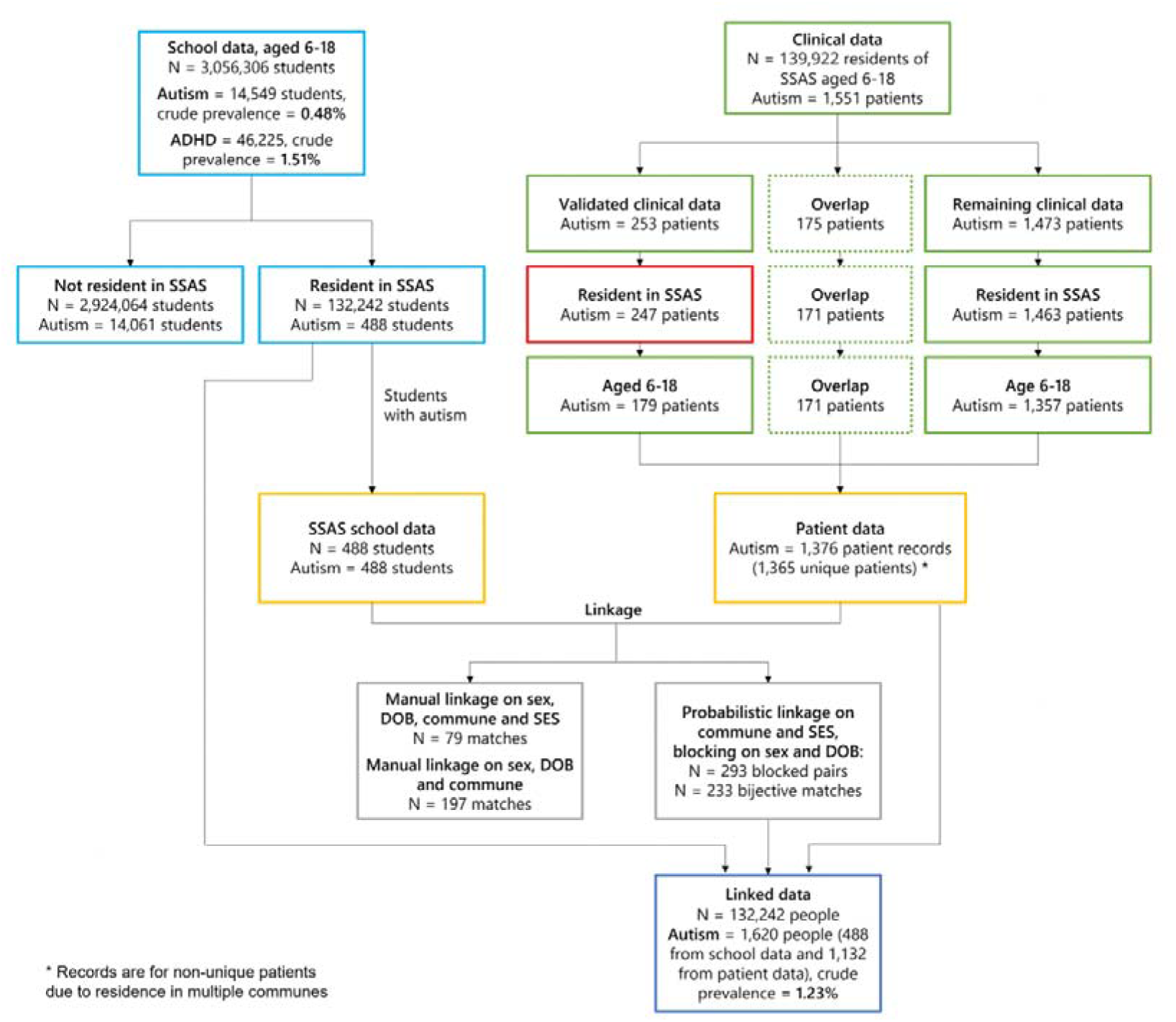
Data Flow of School Data and Clinical Validation Sample.

**Figure 3.**
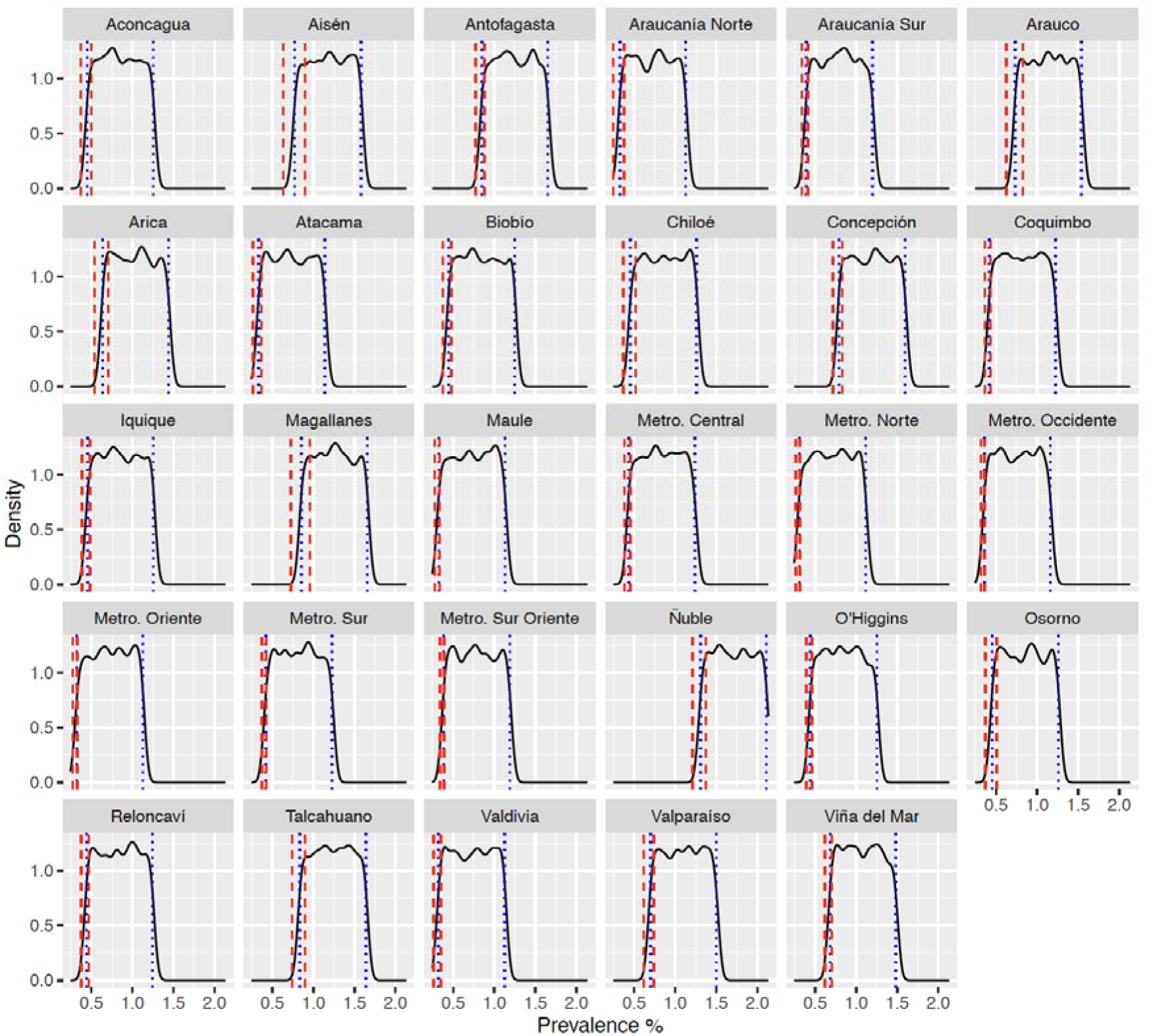
Posterior predictive distribution for health service specific uniform priors. Posterior predictive distributions for ASD prevalence using adjusted case counts from the school data with a random effect on student’s health service. Modelling used uniform priors bounded below by health service specific adjusted ASD prevalence from school data, and bounded above by health service specific adjusted updated ASD prevalence from data linkage. Red dashed lines show the adjusted sample prevalence 95% gamma confidence intervals and blue dotted lines show the posterior 95% credible interval.

## 3. Discussion

There have been significant advances in the field of autism epidemiology in recent years, contributing to a deeper understanding of the prevalence, characteristics, and determinants of an autism diagnosis. One notable approach to enhancing the accuracy and comprehensiveness of epidemiological data is linkage school registries and clinical records. To the best of our knowledge, ours is the largest autism prevalence study in Latin America and the Caribbean to date, and one of the largest to link school registry and clinical records in the world.

This study found an adjusted prevalence rate of 0.46% using school registry data and 1.22% using electronic health records data from the SSAS region, with considerable gender disparity, showing a male-to-female ratio of 6:1 and 4:18:1. This ratio from school registry data is considerably higher than the internationally recognised ratio of 4:1^1,2^, and may suggest diagnostic and awareness bias towards males in school settings. Using Bayesian prevalence estimation to reinvestigate national-level prevalence estimates, we estimated an updated national autism prevalence in Chile of 1.31% (95% CrI 1.25%-1.38%). This also showed that only 30.12% accessed SEN support for autism in the PIE program and that as many as 25,903 out of 40,113 (64.57% unmet need) school age children with autism in Chile do not access the national SEN programme, with boys being 50% more represented in the PIE program, further underscoring the need for focused research into the factors driving these variations. This unmet need, although considerable, includes students with autism diagnoses at non-subsidised private schools who are ineligible for SEED but who may receive other school-based support. It also excludes students who receive SEED for another condition, and students who do not need or do not want school-based interventions. Furthermore, in light of the new 2023 autism law in Chile, it is pertinent to examine how this legislation addresses educational disparities faced by individuals with ASD. This legal framework is an attempt to rectify the systemic shortfalls that historically contributed to the underrepresentation of individuals with ASD in educational settings. It emphasizes the importance of optimizing resources to bridge the gap between diagnosed cases and access to appropriate interventions by adding a gender dimension that could improve access of autistic girls to SEN support at school.

The use of clinical records to validate school registry epidemiological data offers several advantages that improve the accuracy of autism prevalence data from school registries alone. Linking across a wide range of socio-demographic variables enables researchers to perform population-level analyses with large datasets that by themselves might present with incomplete information, helping to identify potential disparities in access to services and resources. From a methodological perspective, our study adds to an emerging body of literature that leverages the ability to link large administrative and clinical data to model more accurate burden of disease estimates using Bayesian methods^20–22^, which is particularly suitable for the more accurate modeling of burdens of disease with fragmented health information systems^21^. That said, the study had some limitations. The national projections of our Bayesian prevalence estimation are based on the assumption that the discrepancy between school and clinical records is uniform across all regions in Chile. While this is a valid starting point for the purpose of this methodology, we encourage follow-up analyses that further examine discrepancies between school and clinical data in order to further refine autism prevalence estimates. While our data captures the majority of Chilean pupils aged 6 to 18 years, our findings may not be directly translatable to other countries in the Latin American and Caribbean region given heterogeneity in health and education systems, as well as possible differences in the make-up of health inequalities^23^. Nevertheless, our findings remain particularly relevant for the purpose of health system planning in Chile, especially in light of the 2023 autism law. We suggest that further research should delve into those areas where disparities in SEN services might exist, including insurance status, which is a crucial variable in measuring access to services and which, in countries such as the US, has not been assessed with regard to its impacts on autism prevalence. Efforts to explore the relationship between autism prevalence, sex, and unmet need for SEN should continue, recognizing the complexities involved in accessing diagnostic services and regional variations in health and educational access across Latin America and the Caribbean.

## 4. Methods

### 4.1 The Chilean PIE School Registry and Electronic Health Record Data

The 2021 Chilean school registry, collected by the Ministry of Education, covers Chile’s total student population together with schools that participate in the PIE program (eMethods 1). The smallest administrative subdivision in Chile is called a commune. The country has 346 communes grouped into 56 provinces, which are, in turn, grouped into 16 regions. This study followed the Strengthening the Reporting of Observational Studies in Epidemiology (STROBE) reporting guidelines and was approved by the Ethics Committee (PRE.2023.021) of the Department of Psychology, University of Cambridge and the Servicio de Salud Araucanía Sur Ethics Committee (Folio: 315). In May 2023, with access to anonymised registry data granted by the Ministry of Education, we obtained anonymised health care contacts between 2014 (year of introduction) and 2021 from the electronic health records of the SSAS. From secondary care records and mental health community services we collated the electronic record data of the health visits of patients aged 6–18 with a primary diagnosis of autism for all communes in the SSAS catchment area. Two datasets were derived from this: (i) a clinical dataset and (ii) a clinical validation subsample, which was a subset of the clinical dataset. The clinical validation subsample was taken from the catchment area of Villarrica Hospital, the second-largest SSAS facility. This subsample was used to evaluate the accuracy of autism diagnoses (see Figure 1).

The small clinical dataset was understood to include every 6–18 year old patient with a primary diagnosis of autism resident in the municipalities of Curarrehue, Loncoche, Pucón and Villarica in the SSAS catchment of Chile’s Araucanía region, and including patients resident in the municipalities of Cunco, Freire, Gorbea, Nueva Imperial, Pitrufquén, Temuco, Teodoro Schmidt, and Toltén. It also included patients resident in the municipalities of Algarrobo (Valparaíso San Antonio health service catchment, Valparaíso region), Cabo de Hornos (Magallanes health service catchment, Magallanes y Antártica Chilena region), Diego de Almagro (Atacama health service catchment, Atacama region), Hijuelas (Viña del Mar Quillota health service catchment, Valparaíso region), Machalí (Libertador B. O’Higgins health service catchment, Libertador B. O’Higgins region), Panguipulli (Valdivia health service catchment, Los Ríos region), Pencahue (Maule health service catchment, Maule region), Pica (Iquique health service catchment, Tarapacá region), Quinta Normal (Metropolitano Occidente health service catchment, Metropolitana de Santiago region) and Tocopilla (Antofagasta health service catchment, Antofagasta region).

### 4.2. Operationalising Autism Status From School Registry Data

The PIE is a school-specific learning program comprising 28 different permanent and transitory categories of provision. Diagnoses are made by child and adolescent psychiatrists or pediatric neurologists registered in *Superintendencia de Salud*, Chile’s health services regulator. The Chilean school registry shows whether students have accessed the PIE through a SEED and whether they require adjustments such as specialist schools or small class sizes. The Chilean school registry includes only one binary coded SEN category per pupil, based on whether a child meets the diagnostic criteria for autism (codes F84.0 to F84.9) from the International Statistical Classification of Diseases and Related Health Problems, Tenth and Eleventh Revision (ICD-10 and ICD-11).

### 4.3. Independent Variables and Regional Analysis Units in the School Registry

Using the school registry dataset, we coded our independent variables as follows: (i) age in four different three year bands (primary school: 6–8 and 9–11; secondary school: 12–14 and 15–18 years), (ii) sex (binarily assigned at birth), (iii) immigration status (yes or no), (iv) monthly school fees converted to US Dollars (Free, $1.15–$11.50, $11.51–$28.75, $28.76–$57.51, $57.52–$115.01, > $115.02, missing), (v) ethnicity (Mapuche, Aymara, Other, and No native groups), and (vi) rurality (yes or no). Chile has 15 regions serviced by 29 health services. We mapped students’ addresses to their respective health service catchment area, with missing address data imputed using the student’s school’s commune. We reported autism school prevalence across Chile’s 29 health services as a measure of access to SEN services. We then selected the SSAS as our clinical validation catchment area. This health service provides primary, secondary, and tertiary health care to 21 communes in Cautín province in the Araucanía region (IX) which has a large urban centre in Temuco and a sizeable rural population (32.8%), and it is the third most populated province in the country after Santiago and Concepción. Its large catchment area (N=752,100) represents 4.3% of the total Chilean population (N=17,574,003) ^8^.

In our clinical validation dataset, we also used a proxy for socioeconomic disadvantage based on health service users’ social health insurance status^6,24^. Membership to Chile’s public insurance scheme is used extensively as a proxy for income given that (i) it is linked to wage-deducted contributions and (ii) privately insured users prefer hospitals in the private health care network and rarely attend public clinics^24^. We further used students’ school fee status as a proxy for socio-economic status. In Chile, the government operates a voucher system that benefits around 93% of primary and secondary students, with the remaining 7% attending private institutions that do not receive subsidies. This system finances schools based on student attendance. The educational institutions involved can be public—typically municipally owned—or private. Private schools, whether they operate for profit or not, can receive state support if they allocate at least 15% of their class seats to students deemed “vulnerable,” which is determined by factors such as family income and the educational level of the mother. These schools also gain additional funding for each vulnerable student they admit. Students with free schooling were assigned low SES as families with low SES are entitled to educational rebates. Students paying betyween $1.15-115.02 US Dollars in fees were assigned medium SES and students paying more than $115.02 monthly were assigned high SES. Student ethnicity was mapped to being a member of the Mapuche Indigenous group, being a member of another Chilean Indigenous group, or not being a member of an Indigenous group based on recorded ethnicity, which could take at most one value. Students with ethnicity recorded as ‘no registry’ were mapped to not being a member of an Indigenous group.

### 4.4. Linking School Registry and Clinical Data

To assess the unmet need for autism in the school registry and its relationship to clinical diagnoses, and to provide corrected national estimates, we used a clinical validation sample. To demonstrate the validity of the clinical diagnoses taken from the electronic health records, a subset of this clinical sample was manually reviewed by a child and adolescent psychiatrist and by a pediatric neurologist, both of whom participate in the PIE diagnostic process. Interrater agreement was computed using Cohen’s Kappa^25^. If a child’s record indicated that the autism case definition had been met, information from the child’s developmental evaluations, PIE plans, and other documents (e.g., cognitive or IQ tests) were obtained and records across data sources were combined. A child met the autism case definition if they were between 6 and 18 years old in 2021, lived in the SSAS health service catchment area during 2021, and had ever received any of the following: a written statement from a qualified professional diagnosing autim, a PIE classification of autism, or ICD-10 codes F84.0 to F84.9. The remaining records were then matched to the Chilean national school registry, using Fellegi-Sunter probabilistic data linkage^26^, based on date of birth, sex, an SES proxy based on insurance status and school fees, and commune of residence. We performed a clerical review of machine-generated matched pairs based on the calculated individual field-similarity scores for each input field, and then reviewed these pairsusing a composite pair-similarity score. The candidate with the highest pair-similarity score was chosen as a match.

For data linkage and to maximise comparability with the clinical data, school registry data were restricted to students with autism that were living in municipalities in the SSAS catchment in 2021. One fabricated empty record was added to the school dataset before linkage to allow the algorithm to correctly match on SES. This fabricated record was only used during linkage, did not match to any patient records, and was removed before matched and unmatched records were compared. This was restricted to appointments for individuals resident in commune in the SSAS catchment as the data for this catchment area are believed to be complete. It was also restricted to patients aged 6–18 years as of 30 June 2021 to maximise compatibility with school data. Appointment year was not restricted in order to retain more data and thus maximise linkage opportunities, and only patients of female and male sex were included.

Sex, date of birth, municipality of residence, autism diagnosis, and the proxies for socio-economic status (i.e., monthly school fees and health insurance contributions mode) were available to match in both school and clinical datasets. Although only students diagnosed with autism are included in the school dataset, the clinical dataset comprised primarily patients diagnosed with autism, but also included some patients diagnosed with an intellectual disability and some diagnosed with both. The autism diagnosis feature was therefore included to encourage matching of school records to clinical records for patients with any type autism diagnosis, as well as to allow matching to clinical records for patients with only a diagnosis of intellectual disability when no suitable patient with autism was present.

All possible pairs of blocked matches were generated and agreement weights were calculated for each feature using expectation maximisation. These feature weights were then aggregated into a weight for each pair. This linkage method is robust to missingness, so observations with missing values were retained. As a similarity comparison method, we used exact matching for municipality of residence, autism diagnosis, and socio-economic status. There was no value in using a string comparison method for municipality of residence as all municipality names were already standardised and two municipalities with similarly spelled names did not increase the likelihood of a match between those municipalities.

To link the datasets, we applied a comparator cut-off value of 0.99 for municipality of residence and autism diagnosis as these are expected to be fairly accurate features. We applied a cut-off of 0.60 for socio-economic status as it is a loosely defined proxy. These values were chosen iteratively through trial-and-error to ensure the algorithm prioritised matching on autism diagnosis above matching on socio-economic status. Linkage was implemented using R’s RecordLinkage package. This included consideration of the average frequencies of categories in each feature and estimated error rates were supplied. The default estimated error rate of 0.01 was supplied for the commune of residence and diagnosis with autism features as these were expected to be fairly accurate features. An estimated error rate of 0.1 was supplied for the socio-economic status feature to reflect that it was a loosely defined proxy. Pairs were then selected based on weight to create a 1-1 bipartite matching between school records and patients. These matches were examined to ensure a patient who lived in multiple municipalities matched to only one school record and to assess the plausibility of matches made to patients diagnosed with intellectual disability instead of autism. Clinical data were subsequently de-duplicated to a single row per patient. In the case of a patient with multiple entries, the matched record for municipality was chosen. School data for students aged 6–18 in the SSAS catchment area and deduplicated patient-level data for patients aged 6–18 in SSAS were combined to form the linked dataset, omitting deduplicated patient records that were a match to student records to ensure such individuals were not present twice in the linked data.

For the school and clinical datasets, each record was classified as either matched or unmatched based on whether it appeared in the bipartite matching. The discrete Kolmogorov-Smirnov test was used to compare matched and unmatched records within each dataset for each of the matching features, with the exclusion of date of birth and autism diagnosis; the former had too many categories to have meaningful results and the latter was uniformly true in the school dataset and therefore not informative. Missing values in the socio-economic status feature were omitted before testing. Permutation tests were then performed for each of the features tested in each dataset by permuting the matched status 2000 times and recomputing the discrete Kolmogorov-Smirnov test for each permutation. The p-values for the Kolmogorov-Smirnov tests on the observed data were then compared to the distributions of p-values for the permuted data to determine the significance of the observed results. For each patient that had lived in more than one commune and therefore appeared more than once in the patient data, only one match to an SSAS school record was made, meaning the matching was bijective for SSAS school records and unique patients.

Kolmogorov-Smirnov permutation tests found no significant difference in frequency of sexes between matched (12.88% female) and unmatched SSAS school records (12.16% female). However, we found a strongly significant difference in the frequency of sexes between matched (12.88% female) and unmatched patient records (21.52% female). This difference was likely due to the difference in male to female ratios across the datasets. SSAS school data for students with autism were 12.47% female, the SSAS patient data were 20.06% female, and matches were 12.50% females. Permutation testing found that matched (39.91% resident in Temuco) and unmatched records (56.08% resident in Temuco) for SSAS school data differed significantly by commune; however, there was no significant difference in the patient data by commune between matched (40.34% resident in Temuco) and unmatched records (40.16% resident in Temuco). This appeared to be driven by the matchability of students and patients living in Temuco, the most populous commune.

### 4.5. Statistical Analyses

#### 4.5.1. Frequentist analysis of school registry data and determinants of access to AUTISM SEN

Data were analyzed from 1 May 2023 to 25 October 2023 in R version 4.3.1^27^. Raw national prevalence estimates for autism in Chilean schools were directly standardized and stratified by age and sex, using the Chilean 2017 census projections for 2021 as the standard population to calculate national prevalence across the country’s 29 health services^2^. To assess determinants to autism SEN access, adjusted adjusted prevalence ratio estimates were obtained using a Poisson regression with robust error variance used to model count data and using autism SEN status as the outcome variable^2^. In each outcome model, we used the same independent variables of sex, age band, immigration status, ethnic group, school fees, and rurality, and included these in the same adjusted model comparing all levels against each other and reporting for missing data. Confidence intervals were calculated at the 95% level using chi-squared distributions. To control for multiple comparisons, we used a significance level of 2-sided PD<0D.001 for all reported outcomes. We then conducted a sensitivity analysis comparing the Poisson regression model to a two-level mixed-effects logistic regression model with two random intercepts at the school and commune level to calculate odds ratios (ORs) for AUTISM SEN access in Chilean schools, adjusting for the same independent variables as in the Poisson model, and comparing model fit.

#### 4.5.2. Bayesian prevalence estimates using school, clinical, and linked data and unmet need for SEN

After calculating the prevalence of autism using school data, we used the linked data to calculate clinical autism prevalence in the SSAS. The adjusted prevalence delta, was calculated as the ratio difference between the adjusted prevalences for the (a) SSAS linked data and (b) school registry data. For the purpose of this analysis, we assumed this ratio was applicable nationally. This ratio was then extrapolated to the country’s other 28 health services using SSAS as prior in a Bayesian random-effects health service model to calculate the adjusted prevalence projections. Estimated credible intervals were calculated for the projections by finding the maximum band around the projection of equal width to the 95% gamma confidence interval for the adjusted prevalence of the school data for each health service. Using Bayesian prevalence analysis of autism to calculate autism prevalence inference with different types of incomplete data allowed plausible national prevalence estimates and provided information about the likelihood of these predictions given the observed school data. We used the following

Bayesian model:

(1) y_i_|(n_i_, θ_i_) ∼ Binomial(n_i_, θ_i_)

in which y_i_ refers to the adjusted count of autism cases in health service i, n_i_ represents the number of students in health service i, and θ_i_ designates the prevalence of autism in health service i. This model is paired with a prior distribution of θ_i_ that follows:

(2) θ_i_ ∼ Beta(a, b) in which a captures a prior of the autism prevalence rate and b represents the corresponding prior of the standard deviation of the autism prevalence rate. When combined, the subsequent posterior distribution can be derived to model the autism prevalence rate in health service i given the number of children diagnosed with autism in health service i and the total population of health service i:

(3) θ_i_|(y_i_, n_i_) ∼ Beta(y_i_ + a, n_i_ − y_i_ + b)

Because we had complete data on autism in the school registry as well as clinical records from one regional health service, we used four priors for θ_i_ when fitting the Bayesian prevalence model to account for data imbalances at two different levels (a) national at the (b) health service level and for lower and upper bounds of prevalence in (c) schools and in (d) clinical population. To calculate a national prevalence with the clinical and school registry information, we had and then a health service prevalence rate under the assumptions we obtained from our data linkage.

We used a (i) conjugate beta prior common to all health services as our first prior, constructed with the national adjusted autism prevalence from the school registry and its standard deviation as the mean and standard deviation of the prior. This prior was suitable because the adjusted prevalence in the school data provides a plausible lower bound on the prevalence of autism in Chile. This lower bound is a general starting point for the prevalence of autism across all health services and acts as a plausible lower bound for autism prevalence, providing a conservative estimate that informs the initial baseline for all regions before more specific data are considered.

We used a (ii) health service-specific conjugate beta prior, developed using the health service-specific adjusted autism prevalence estimates from the school registry and their respective prior means and standard deviations. This prior was also suitable because it extended the previous prior to each of the random effect categories and reflected students receiving SEN. On its own, this prior was expected to give uninformative posteriors because it effectively duplicated the information in the sample data. However, it was suitable as a more specific lower bound on the plausible prevalence of autism in each health service.

The third prior was a (iii) conjugate beta prior based on linked data, which is specific to each health service, derived using the adjusted prevalence projected from the linked data from SSAS electronic health records and their standard deviations from their maximal 95% confidence intervals as the prior means and prior standard deviations respectively. This prior was suitable as it captured the clinical information provided by the linkage and included all students with clinical diagnoses and or with autism SEN recorded in the school registry; additionally, it had narrow standard deviations which modeled a theoretical upper bound on the prevalence of autism in each health service.

Our fourth prior was a (iv) uniform prior specific to each health service, created using the adjusted autism prevalences from the school data for each health service as its lower bounds, and the projected prevalences ratio from the linked data for each health service as its upper bounds. This prior was suitable because it captured the information from both the school and linked datasets, without specifying where within these bounds the true prevalences were likely to be.

Bayesian prevalence modelling was implemented in the Just Another Gibbs Sampler (JAGS) language, which uses Markov chain Monte Carlo (MCMC) sampling to produce posterior density distributions when given the above priors and adjusted prevalence observations^28^. A burn-in period of 2000 samples was used to ensure models converge. Finally, 2000 iterations without thinning were used to model the posterior densities while checking for convergence^29^.

We employed a novel methodological framework to estimate the prevalence of autism in Chile, utilizing a combination of school registry and clinical health record data. The integration of these datasets through probabilistic data linkage techniques allowed for the analysis of both diagnosed cases and access to Special Educational Needs (SEN) services. By employing both frequentist and Bayesian statistical methods, we calculated direct standardized prevalence rates and adjusted estimates that incorporate regional variations and the completeness of data. Specifically, the Bayesian approach utilized four distinct types of priors, including conjugate beta priors and uniform priors, which facilitated a nuanced estimation of autism prevalence across different health services and nationally. These methods not only confirmed the validity of the autism diagnoses through clinical validation subsamples but also highlighted significant regional disparities in the availability of SEN resources. The findings of this study are instrumental for policymakers and educational authorities, providing them with detailed insights necessary for targeting interventions and improving the educational and health outcomes for children with autism throughout Chile.

## Data Availability

Data is not available due to privacy concerns

